# A randomized trial of index HIV self-testing for sexual partners of ART clients in Malawi

**DOI:** 10.1101/2022.09.28.22280455

**Authors:** Kathryn L Dovel, Kelvin Balakasi, Khumbo Phiri, Frackson Shaba, Ogechukwu Agatha Offorjebe, Sundeep K Gupta, Vincent Wong, Eric Lungu, Brooke E Nichols, Mike Nyirenda, Tobias Masina, Anteneh Worku, Risa Hoffman

## Abstract

**Background:** HIV testing among the sexual partners of HIV-positive clients is critical for case identification and reduced transmission. Current strategies have limited reach. We evaluated an index HIV self-testing (HIVST) intervention among ART clients in Malawi, whereby clients were asked to distribute HIVST kits to their primary sexual partners.

**Methods:** We conducted an individually randomized, unblinded trial at 3 district hospitals in Malawi between March 28 2018 – January 5, 2020. Clients attending ART clinics were randomized 1:2·5 to: (1) standard partner referral slip (PRS); or (2) index HIVST. Inclusion criteria were: ART client is ≥15 years of age; primary partner with unknown HIV status; no history of interpersonal violence with that partner; and partner lives in facility catchment area. The primary outcome was completion of index partner testing, and, if positive, index partner ART initiation within 12-months. Baseline and follow-up surveys with ART clients measured the primary outcome and medical chart reviews measured ART initiation. Uni- and multivariate logistic regressions were conducted.

**Findings:** A total of 4,043 ART clients were screened and 456 were eligible and enrolled. 365 completed a follow-up survey and were included in the final analysis (22% men). Testing coverage among partners was 71% in the HIVST arm and 25% in PRS (AOR:9·6; 95% CI: 6·45-12·82). HIV positivity rates did not differ by arm (19% in HIVST versus 16% in PRS; p=0·74). ART initiation at 12-months was 46% (14/30) in HIVST versus 75% (3/4) in PRS arms; however, HIVST still resulted in a 94% increase in the proportion of all partners initiating ART due to high testing rates. Adverse events did not vary by arm.

**Interpretation:** Index HIVST significantly increased HIV testing and ART initiation among ART clients’ sexual partners without increased risk of adverse events. Additional research is needed to understand and improve ART initiation within index HIVST.

**Funding:** United States Agency for International Development under cooperative agreement AID-OAA-A-15-00070. KD receives funding from Fogarty International Center K01-TW011484-01.

**Research in Context:** *Evidence before this study:* Index partner testing, whereby partners of inidivuals living with HIV are tested for HIV, is a primary entry point to HIV services among higher risk populations in eastern and southern Africa. Yet coverage for index partner testing remains poor. Distance to facilities, fear of unwanted disclosure and lack of privacy, and logistics related to tracing partners in the community are all major barriers to uptake of index partner testing. HIV self-testing is an effective strategy to improve testing coverage, but it has rarely been used in the context of index partner testing. HIVST may allow partners to test where and when they want, and may encourage positive communication within partnerships.

*Added value of this study:* We present new evidence from a cluster randomized control trial in Malawi that index HIVST among the primary partners of ART clients can dramatically increase uptake of index partner testing, with a 167% increase in testing compared to standard partner referral slips. Very few adverse events were reported in either arm. We also present some of the first data on time to ART initiation after a reactive HIVST kit, and the cost-effectiveness of an HIVST intervention for ART initiation.

*Implications of all the available evidence:* Index HIVST can increase HIV testing among partners of ART clients without increasing adverse events in Malawi. Importantly, we found that male partenrs were still less likely than female partners to test and initate treatment within the HIVST intervention. Additional interventions to improve linkage to care after using HIVST kits are needed. Index HIVST can be a useful strategy to easily increase testing coverage among higher risk parnters. However, we found that only 9% of ART clients screened had partners who were eligible for index HIVST. This suggests that while index HIVST is effective in the Malawi setting, the intervention’s reach at a national level may be narrow.

## Introduction

Index partner testing, whereby the sexual partners of individuals living with HIV are tested for HIV, is critical to reaching the first 95 of the UNAIDS 95-95-95 goals.^1^ Index partner testing is associated with higher testing yield compared to other case-finding strategies^2-4^ and high rates of status disclosure among partners, which can promote ART adherence.^5-7^ Index partner testing is also critical for identifying individuals who are HIV-negative and can benefit from HIV prevention measures.^8-10^ However, current index testing strategies have limited reach. In Malawi, standard of care referrals (usually paper forms given for partners) result in only 22% of partners tested for HIV.^11^ Male partners are least likely to test. Voluntary assisted partner disclosure and testing, whereby health care workers (HCWs) visit index partners in the community to provide HIV testing services, promises to increase testing uptake^12^, but is costly^13^ and has increased risk for coercion and unwanted disclosure for the ART client, especially among women living with HIV.^14-17^ New strategies are needed to effectively reach partners with index testing, particularly men.

HIV self-testing (HIVST) offers an alternative strategy to improving index partner testing by allowing people living with HIV to take an HIVST kit home to their sexual partner (“index HIVST”). HIVST can be used in the privacy of partners’ own homes and at times convenient to them,^8^ and can largely address barriers to standard index partner testing, including distance to the health facility, time associated with seeking HIV testing, and concerns related to privacy of HIV testing and unwanted disclosure.^18^ Oral-based HIVST has been widely used throughout sub-Saharan Africa and has been shown to be highly acceptable among hard-to-reach populations, including men.^19,20^

A growing body of literature shows that women seeking antenatal (ANC) services and female sex workers are willing and able to distribute HIVST to their male partners,^21-23^ with 79-91% of male partners reported as using an HIVST kit with minimal adverse events.^24^ However, no study to our knowledge has examined the impact of secondary distribution of HIVST kits for partners of individuals living with HIV. Acceptability and feasibility of index HIVST may differ substantially among individuals living with HIV, as ART clients may fear unwanted status disclosure to their sexual partner and may be at higher risk of adverse events, such as interpersonal violence (IPV) and/or the ending of a relationship.

Further, the barriers to and frequency of partner confirmatory testing and ART initiation after receiving a positive HIVST result at home are still unclear.^18^ Reported ART initiation rates among HIVST users vary between 23-68%.^25-28^ Partners of individuals living with HIV may be more likely to initiate ART as compared to the general population given that they may already be exposed to ART services (if their partner is actively in care) and have immediate social support for navigating ART clinics, although this has not been examined to our knowledge.

We conducted an individually randomized controlled trial (RCT) in Malawi to assess the impact of index HIVST on testing uptake among ART clients’ primary sexual partners as compared to standard of care PRS, and describe ART initiation among newly diagnosed individuals.

## Methods

We conducted an individually randomized, unblinded trial among ART clients and their partners in Malawi between March 28 2018 – January 5 2020. The primary outcome was partner status ascertainment 4 weeks after enrollment. Secondary outcomes included: testing yield, usability and acceptability of index partner HIVST, and presence of adverse events, all from ART client secondary reports, and ART initiation rates at 12-months, obtained through medical chart reviews.

### Study Sites and Setting

A convenience sample of three high-burden referral hospitals in central and southern Malawi were included in the trial. At the time of the study, the national HIV prevalence was 9.6%^29^, and 36-49% of adults in Malawi had been tested for HIV in the prior year, with Malawi national guidelines stipulating that index testing should be offered to all partners of clients living with HIV.^30^

### Study Population

Individuals living with HIV and on ART were recruited during routine ART clinic visits. Eligibility criteria included: ≥15 years of age; have a sexual partner in the last 12 months with an unknown HIV status, defined as never tested HIV-positive or not tested HIV-negative within the past 3-months (if multiple partners, individuals were asked to define the HIV status of their primary partner); no history of IPV with that partner; and partner lives within the facility catchment area.

### Randomization and Masking

Participants were randomized 1:2·5 to one of two arms: (1) Standard of care partner referral slip distribution (PRS), whereby ART clients were counseled on the importance of index HIV testing and strategies for disclosure, and offered a PRS slip and linkage card with a map of the facility to give to their primary partner; (2) HIVST, whereby ART clients were counseled on the importance of index HIV testing and strategies for disclosure, given a 10-minute demonstration on how to use the HIVST kit, given one Oraquick oral HIVST kit © for their primary sexual partner, an instructional leaflet on how to use HIVST, and a linkage card with a map to the health facility. To allow for the possibility that partners may be uncomfortable with HIVST, ART clients in the HIVST arm were also given a PRS so that a partner could opt out of HIVST and test via standard HIV testing methods at the nearest health facility. Clients who were confirmed HIV-positive were referred to the facility’s ART clinic for same-day ART initiation.

### Study Procedures

ART clients were recruited by study staff while waiting for routine ART services. Individuals who consented for screening were taken to a private space within the health facility to complete screening, and, if eligible, provide written informed consent. Computer-generated randomization was used to assign clients to either the PRS or HIVST arms in a ratio of 1:2·5, respectively. Immediately following randomization, the intervention (PRS or index HIVST) was delivered by a trained study staff member.

Following counseling and delivery of PRS or HIVST, ART clients completed a baseline survey to understand: (1) socio-demographics of the ART client and partner; (2) history of HIV service utilization; and (3) relationship characteristics with the index partner. ART clients were scheduled for a follow-up 4 weeks after the initial study visit at the same health facility in order to assess primary and secondary outcomes. Follow-up surveys included the following domains: (1) distribution and acceptability of the intervention (PRS or HIVST); (2) index partners use of HIV services since enrolment; and (3) any adverse events experienced since enrolment, such as interpersonal violence (IPV) or termination of the sexual relationship. Both baseline and follow-up surveys lasted approximately 60 minutes and were conducted in the local language (Chichewa) by trained study staff. Identifiers were collected for index partners who were reported to have a positive HIV test during the study period (i.e., name, age, address, ART number – if any). Study staff conducted medical chart reviews at all PEPFAR-supported facilities in the study districts at 3-, 6-, and 12-months post-enrollment to assess if index partners initiated ART. At the end of 12-months, those with positive HIV tests who were not found in medical chart reviews were contacted by facility staff (via phone or in person) in order to ascertain ART initiation outcomes, per standard of care facility protocols.

In the HIVST arm, we also recruited a subset of index partners to complete one follow-up survey. ART clients in the HIVST arm were given a study recruitment card requesting their partner present at the health facility for a study survey approximately 4 weeks after the ART client enrolled in the study. Index partners were surveyed if they were ≥15 years of age and presented at the health facility in order to complete a survey. Screening, written consent, and the survey lasted approximately one hour and were conducted in the local language (Chichewa) by a trained study staff member in a private space. The partner survey was used to assess partner acceptability of index HIVST and ability to use and interpret HIVST.

### Study Outcomes

The protocol-defined primary outcome was the proportion of index partners who tested for HIV within 4 weeks after enrollment of the index client (including either HIVST or standard blood-based testing) and was measured by secondary report from the ART client.

Secondary outcomes included: (1) proportion of ART clients who reported distributing the intervention to their partner; (2) HIV positivity rate among partners tested; (3) ART initiation within 12-months after receiving a reactive blood- or self-test (using medical charts); (4) acceptability and usability of the intervention (PRS or HIVST) from the perspective of the ART client (variables measured on a 4-point Likert scale and dichotomized for analysis as agree/strongly agree or disagree/strongly disagree); and (5) presence of adverse events for the ART client, such as IPV (measured using questions adapted from the Malawi Demographic and Health Survey^32^), including emotional or verbal violence or the end of a relationship the study period.

Total cost per individual testing positive and per individual linked to care were calculated using study data for both study arms. Costs were derived from a recent facility-based HIV self-testing trial.^33^ For the PRS arm, the costs included: cost of counseling when given the PRS, cost of the partner testing at the clinic for those who returned, and the cost of confirmatory testing for those who tested positive. For the index HIVST arm, the costs included: cost of counseling when the HIVST kits were distributed, the cost of the distributed self-tests, and the cost of the clinic-based testing algorithm following a positive HIVST. Costs of human resources, equipment and overhead were adjusted from 2017USD to 2018USD to reflect the time period of this trial. Cost of HIV testing kits (both HIVST and provider testing kits) did not change between 2017 and 2018. The cost of an HIVST kit was assumed to be $2.

### Statistical Analysis

We used the CONSORT standards for reporting trial outcomes. All analyses were prespecified based on the protocol (see Appendix A). Descriptive statistics (mean, standard deviation, median, inter-quartile range, and frequency distribution) were generated for all demographic and clinical information to characterize the study population overall and by study arm. ART clients who did not complete a four-week follow-up survey were excluded from the analyses because it was impossible to determine primary and secondary outcomes. Among those retained, anyone who had missing data or were unsure of primary outcomes were counted as failures (i.e. no HIV test completed). Individuals where an outcome was not ascertained (for example, ART client did not know if their partner tested for HIV) were treated as index partner testing failures. Univariate and multivariate logistical regressions were used to compare primary outcomes between arms. We used random effects to account for correlations within study sites. Secondary outcomes were described using descriptive statistics and were explored using univariate and multivariate logistical regressions, where possible.

Three types of cost outcomes were assessed: 1) Cost per test provided: calculated as the cost per person counseled and given PRS or HIVST. 2) Cost per person aware of status: calculated as the total cost of providing PRS or HIVST to all individuals in the trial divided by the number of people newly aware of their HIV positive status (by arm). 3) Cost per ART initiate: calculated as the total cost of providing PRS or HIVST to all individuals in the trial divided by the number of people newly initiating on ART (by arm).

### Ethics Statement and Participants’ Safety

Participants provided written consent and were given 2·5USD for each study visit. The study was approved by the Malawi National Health Sciences Research Committee (NHSRC) and Institutional Review Board at the University of California Los Angeles (UCLA) USA. The trial is registered with ClinicalTrials.gov, NCT03271307, and Pan African Clinical Trials, PACTR201711002697316.

## Results

A total of 4,043 ART clients (1,688 men and 2,355 women) were screened. Figure 1 shows the recruitment and enrollment of ART clients. Of those approached for screening, 18 declined to participate and 3,679 (91%) did not meet eligibility criteria. The primary reasons for exclusion were partner had a known HIV status (53%) and ART client did not have a sexual partner at the time of recruitment (30%). A total of 130 ART clients were randomized to the PRS arm and 326 to the HIVST arm. Of that, 365 ART clients completed a follow-up survey 4 weeks after enrollment, with similar retention rates by arm (∼83%; 107 in PRS and 258 in HIVST). Primary outcome results are reported for the 365 ART clients (with secondary reports for their partners).

**Figure 1:**
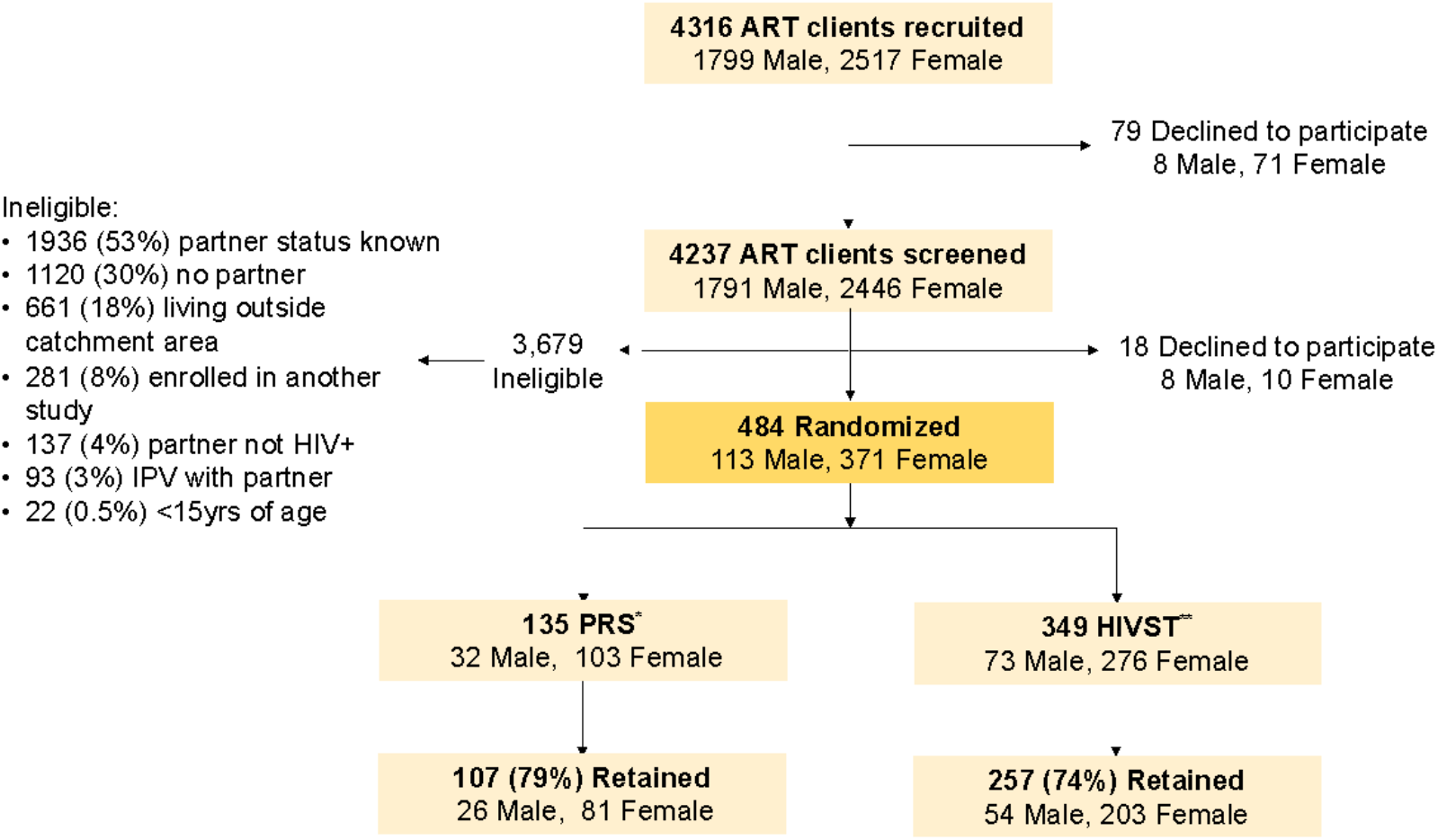
Recruitment and Enrollment of ART clients. *Partner referral slip, ** HIV self-testing

Table 1 shows characteristics of ART clients and their index partners among those for whom a primary outcome was available (n=365). Clients had been on ART for a median 4.3 years (IQR 2-8), 78% were female, and the mean age was 37 years (IQR 29-44). Nearly 83% of ART clients were married or living with their index partner, with mean relationship length of 9 years (IQR 2-15) and a mean of 3 (IQR 2-5) living children (62% had at least one living child with their primary partner). The majority of ART clients (91%) had disclosed their HIV status to their index partner prior to enrolling in the trial and talked with this partner at least once per week (96%). Demographics were similar across arms.

**Table 1.** Baseline characteristics, reported by ART client (n=365)

| Respondent Characteristic | PRS Arm<br>n(%) | HIVST Arm<br>n(%) | Total<br>n(%) |
| --- | --- | --- | --- |
| n | 107 | 257 | 364 |
| **Client Information** |  |  |  |
| **Demographic variables** |  |  |  |
| Male | 26 (24%) | 54 (21%) | 80 (22%) |
| Mean age, years (SD) | 37 (12.4) | 37 (11.2) | 37 (11.6) |
| Median years of education, (IQR) | 4 (1-8) | 5 (1-8) | 4 (1-8) |
| Household wealth quantiles |  |  |  |
| Low quantile | 41 (38%) | 81 (32%) | 122 (34%) |
| Middle quantile | 29 (27%) | 92 (36%) | 121 (33%) |
| High quantile | 37 (35%) | 84 (33%) | 121 (33%) |
| Worked for pay in last month (formal or informal) | 55 (51%) | 137 (53%) | 192 (53%) |
| Median number of living children, (IQR) | 3 (2-5) | 3 (2-5) | 3 (2-5) |
| **HIV services** |  |  |  |
| Median years on ART, (IQR) | 4.2 (1-9) | 4.3 (2-7) | 4.3 (1-8) |
| Disclosed their HIV status to a non-sexual partner | 96 (90%) | 228 (89%) | 324 (89%) |
| Have a close friend on ART (talk with them at least 1/week) | 51 (54%) | 119 (55%) | 170 (54%) |
| More than one sexual partner in past 12 months | 10 (9%) | 19 (7%) | 29 (8%) |
| **Primary Partner Information** |  |  |  |
| **Demographic variables** |  |  |  |
| Male | 81 (76%) | 203 (79%) | 284 (78%) |
| Mean age, years (SD) | 40 (12.5) | 40 (11.1) | 40 (11.5) |
| **Relationship with primary partner** |  |  |  |
| Disclosed HIV status to primary partner | 93 (90%) | 232 (92%) | 325 (92%) |
| Median years in a relationship, (IQR) | 4 (2-14) | 6 (2-15) | 5.5 (2-15) |
| Relationship type with primary partner |  |  |  |
| Married/living together | 88 (82%) | 214 (83%) | 302 (83%) |
| Steady partner | 16 (15%) | 28 (11%) | 44 (12%) |
| New/Infrequent partner | 3 (3%) | 15 (6%) | 18 (5%) |
| Talk with partner at least once per week | 98 (95%) | 240 (96%) | 338 (96%) |
| Has children with the primary partner | 63 (61%) | 164 (66%) | 227 (64%) |
| Confident they would still be with this partner in 12 months | 82 (80%) | 209 (84%) | 291 (82%) |
| **Region** |  |  |  |
| Southern | 72 (67%) | 157 (61%) | 229 (63%) |
| Central | 35 (33%) | 100 (39%) | 135 (37%) |

### Primary Outcomes

Reported HIV testing among index partners within 4 weeks of enrollment was significantly higher in the HIVST arm as compared to the PRS arm (71% versus 27%, respectively; AOR: 9·09, 95% CI: 6·45 - 12·82; see Figure 2). Testing coverage increased with index HIVST relative to PRS by 46% in male index partners and by 50% in female index partners (see Table 2).

**Table 2.**
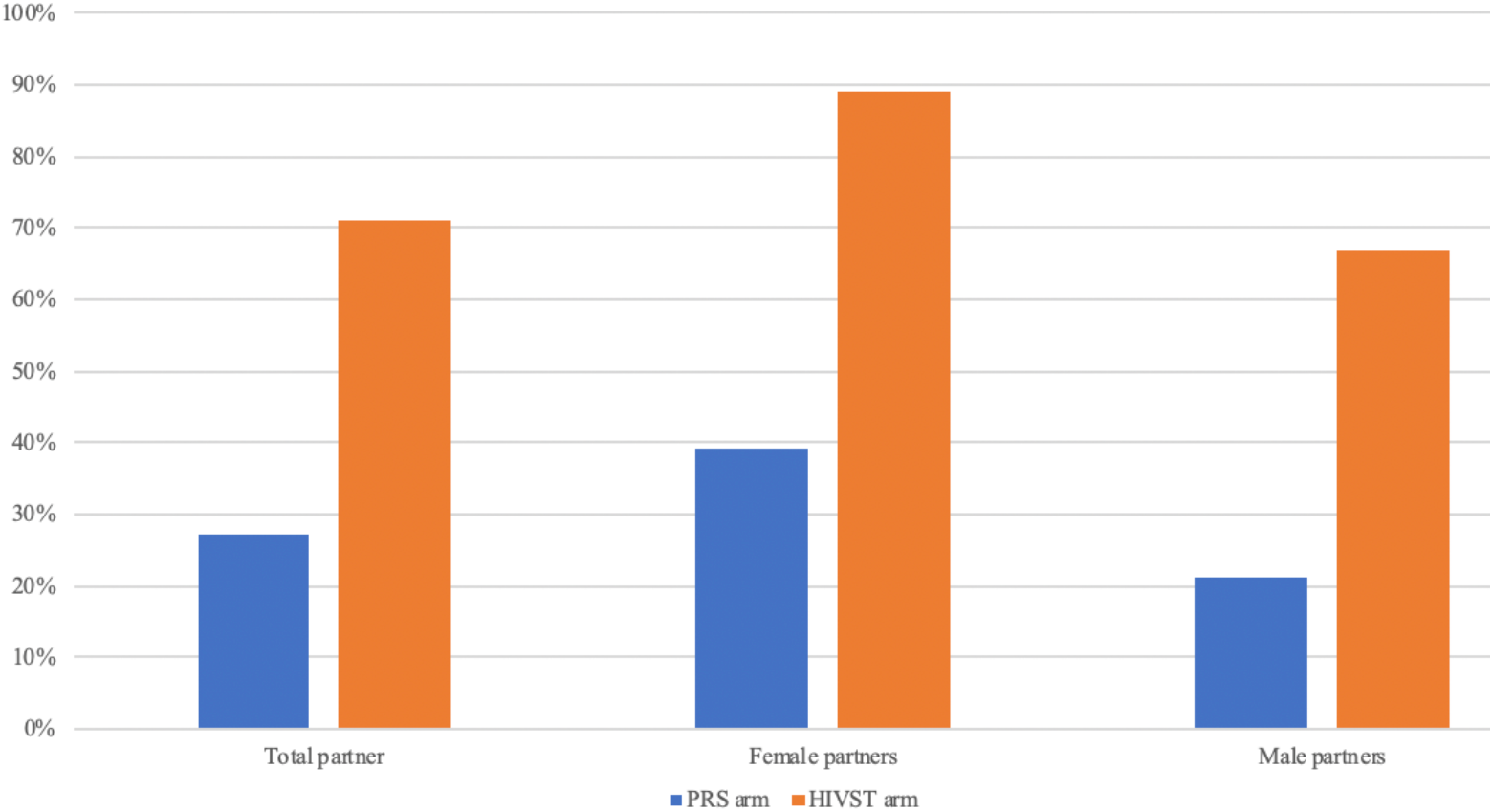
HIV testing among partners of ART clients, by sex across trial arm (n=364)

### Secondary Outcomes

#### Distribution of Intervention to Index Partner

The vast majority of ART clients in both arms reported having distributed the intervention to their index partner (92% for PRS and 91% for self-test; p-value=0·74). Reasons for not delivering either PRS or HIVST were similar across arms: 16/32 (50%) had not seen their partner since enrolling in the study, 5/32 (17%) were afraid of their partner’s response, and 4/32 (13%) were afraid to disclose their status to their partner.

### HIV Positivity Rate

HIV-positivity among index partners who tested was similar across arms, with 15% (4/27) testing positive in the PRS arm and 16% (30/183) in the HIVST arm. While positivity rates were similar across arms (p-value = 0·84), high testing coverage among HIVST partners translated to a greater proportion of partners identified as HIV-positive (30/257; 11%) as compared to PRS (4/107; 4%) (AOR:3·40; p-value=0·03).

### ART Initiation

All index partners in the PRS arm who tested HIV-positive (n=4) did so at a health facility and 3/4 (75%) initiated ART on the same day as testing. In the HIVST arm, 14/30 (47%) of the index partners who tested HIV-positive were identified as having initiated ART within 12 months of their test, the vast majority doing so within 3-months of testing (13/14, 93%; see Appendix B).

**Table 2.** HIV testing uptake among index partners, reported by index (ART) clients.

| HIV Testing Service | PRS Arm<br>n(%) | HIVST Arm<br>n(%) | OR (95% CI) | AOR (95% CI)<sup>^</sup> |
| --- | --- | --- | --- | --- |
| Index client delivered intervention | 98/107 (92%) | 231/257 (90%) | 0.93 (0.28 - 3.12) | 1.10 (0.29 - 4.24) |
| Female index partner | 25/26 (96%) | 52/54 (96%) | 1.04 (0.15 - 7.04) |  |
| Male index partner | 73/81 (90%) | 179/203 (90%) | 0.95 (0.22 - 4.14) |  |
| Index partner tested | 27/107 (25%) | 183/257 (71%) | 7.33*** (5.67 - 9.48) | 9.09*** (6.45 - 12.82) |
| Female index partner | 10/26 (39%) | 48/54 (89%) | 12.80*** (4.35 - 37.65) |  |
| Male index partner | 17/81 (21%) | 135/203 (67%) | 7.47*** (5.72 - 9.77) |  |
| Index partner tested HIV-positive | 4/27 (15%) | 30/183 (16%) | 1.13 (0.37 - 3.46) | 0.99 (0.26 - 3.73) |
| Female index partner | 0/10 (0.0%) | 2/48 (4%) | - |  |
| Male index partner | 4/17 (24%) | 28/135 (21%) |  |  |
| HIV-positive index partner-initiated ART at 12-months | 3/4 (75%) | 14/30 (47%) |  |  |
| Female index partner | 0 (0.0%) | 0/2 (0.0%) |  |  |
| Male index partner | 3/4 (75%) | 14/28 (50%) |  |  |

<sup>^</sup>adjusted for age, sex, and marital status, \* $p < 0.1$ , \*\* $p < 0.05$ , \*\*\* $p < 0.01$

### Acceptability and Adverse Events

Table 3 shows intervention acceptability and adverse events outcomes as reported by ART clients. Over 98% of ART clients in both arms were comfortable explaining the respective intervention, and 99% of ART clients in the index HIVST arm were comfortable demonstrating HIVST to their index partner.

**Table 3.**
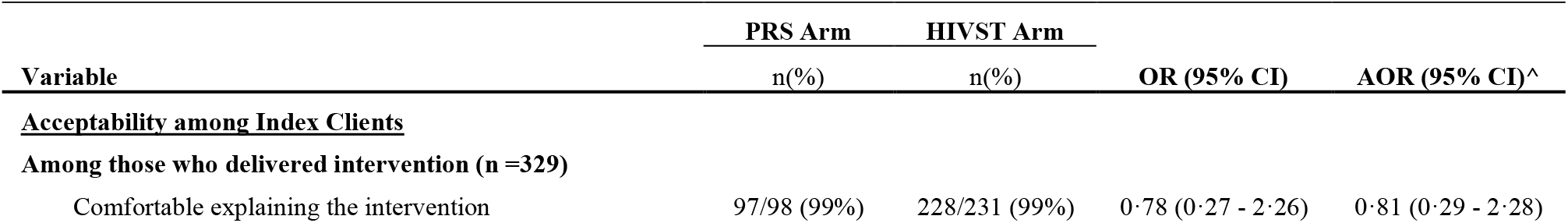
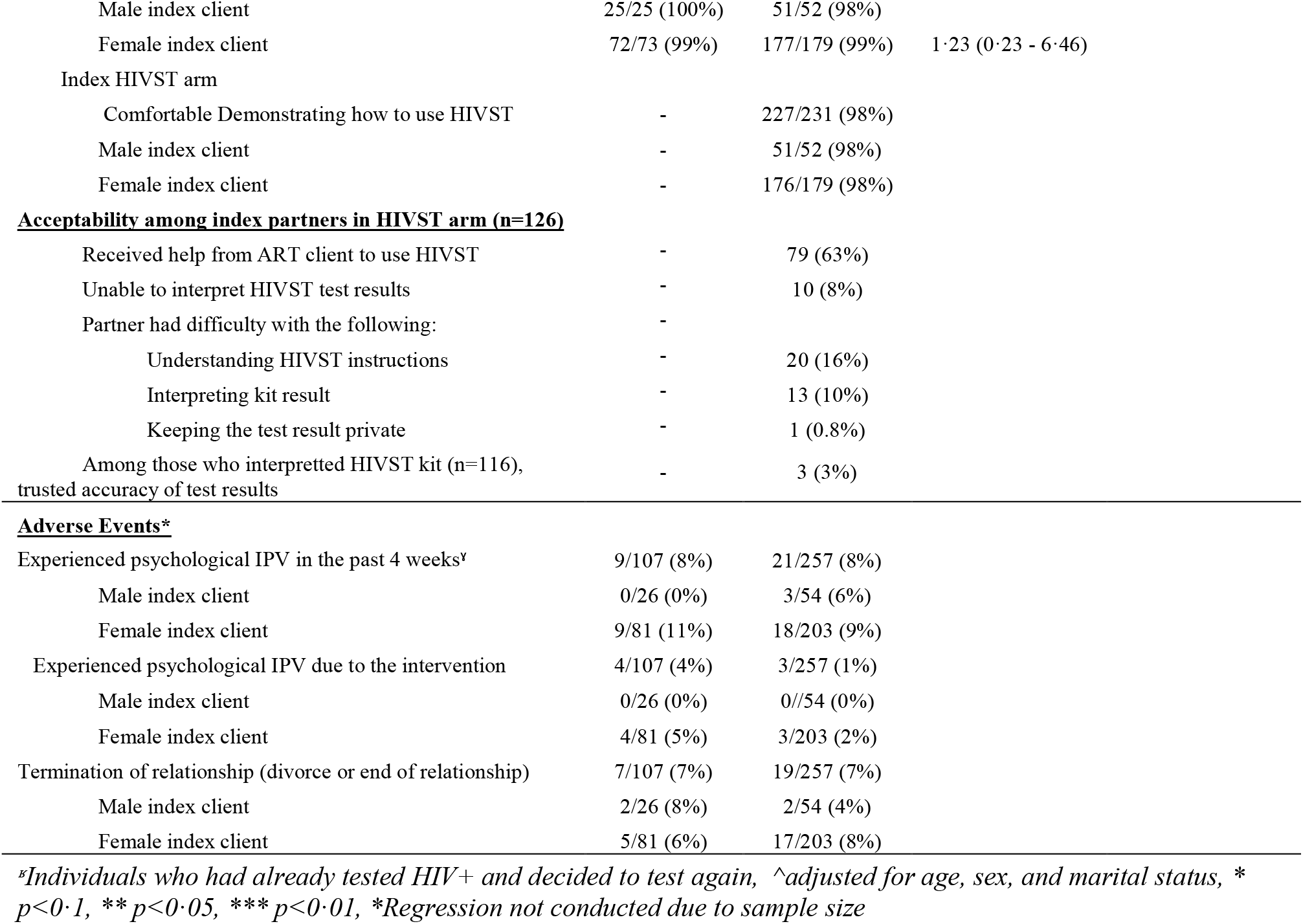
Acceptability of the intervention and adverse events, reported by Index clients (n=364)

**Table 4.** HIV testing uptake among index partners, reported by index (ART) clients (n=364)

| Costing | PRS Arm | HIVST Arm |
| --- | --- | --- |
| **Costing inputs** |  |  |
| Cost of counselling for PRS or index HIVST distribution* | \$0.15 | \$0.23 |
| Cost per HIVST kit | - | \$2.00 |
| Cost of testing negative at health care facility** | \$2.29 | \$2.29 |
| Cost of testing positive at health care facility** | \$3.85 | \$3.85 |
| **Costing outcomes** |  |  |
| Cost per test provided | \$0.84 | \$2.34 |
| Cost per person aware of HIV status | \$19.35 | \$16.06 |
| Cost per ART initiate | \$24.46 | \$84.15 |

\*All inclusive costs (human resources, equipment, overhead)
\*\*All-inclusive costs (human resources, equipment, test kits, overhead)

We also conducted a survey with a subset of index partners in the HIVST arm who attempted to use HIVST (n=126). Mean age of surveyed partners was 41 (IQR: 34 - 46) and 73% were male (see Appendix B for demographic details). Among these index partners, 16% reported challenges understanding HIVST instructions and 8% were unable to interpret HIVST results. Among those able to interpret their test results (n=116), only 3% did not trust test results (and all tested HIV-negative).

Adverse events as reported by ART clients did not vary by arm. Eight percent of ART clients across both arms reported psychological IPV in the 4-weeks after study enrolment. Four-percent of those in the PRS arm and 1% in the HIVST arm reported psychological IPV events that they believe were due to the assigned intervention arm (*p*-value=0.57). A ll ART clients who reported IPV were female. There were no reports of physical or sexual IPV. Termination of relationship status was 7% in both arms (p-value=0.89).

The cost per test provided was lower for PRS ($0·84) than for HIVST ($2·34). This is explained by the fact that not everyone who received a PRS received a test, whereas every HIVST distributed incurs the cost of an HIVST kit. The cost per person newly aware of their positive status was lower for HIVST than for PRS ($16·06 and $19·35 respectively) given that more people used the self-test kit than followed up using their partner referral slip. However, the cost per new ART initiate was far greater for HIVST as compared to PRS ($84·15 vs $24·46). This is driven by individuals who tested positive using HIVST but did not follow-up at a health care facility for a confirmatory test and ART initiation.

## Discussion

In this RCT, we found that index HIVST among the primary partners of ART clients dramatically increased the proportion of partners tested for HIV. The index HIVST arm showed a 167% increase in testing uptake as compared to standard partner referral slips (PRS), achieving testing coverage of 71% among sexual partners. Due to increased testing coverage, index HIVST resulted in a 211% increase in the number of partners identified as living with HIV as compared to PRS. After 12-months, 14/30 (47%) of those newly diagnosed in the HIVST arm and 3/4 (75%) of those in the PSR arm initiated ART. However, the HIVST arm also showed nearly double the proportion of total partners initiating ART – even with lower initiation rates – due to dramatic increases in testing coverage (5·4% of the total HIVST sample initiating ART versus 2·8% of the total PRS sample). Findings suggested that index HIVST can effectively increase testing uptake and ART coverage among partners of ART clients, although additional work is needed on linkage to fully understand differences between PRS and HIVST.

The intervention was cost-effective for new diagnoses, and the cost per newly diagnosed individual was low compared to other testing modalities that target general populations (such as community screening, facility based HIVST, among others).^34^ This is likely due to the high probability of index partners being positive as compared to people tested through other modalities. However, we found that only 9% of ART clients screened had partners who were eligible for index HIVST. This suggests that while index HIVST is effective in the Malawi setting, the intervention’s reach at a national level may be narrow and limit the overall impact of index HIVST for national programs.

A sub-analysis of our data showed large testing differences by sex – in the HIVST arm 89% of female partners tested versus 67% of male partners. This disparity is important as index testing represents a key strategy for reaching men, who comprised over 78% of eligible index partners and, as shown in other literature, may not have other clear entry points into HIV services.^35,36^ A package of strategies may be needed in order to optimally reach male partners, such as index HIVST plus peer support for men or male targeted promotional messaging.^24,37^ In addition, other entry points such as outpatient departments, the only facility-based entry point reached by most men, may need to be utilized further to reach men.

Index HIVST was highly acceptable among ART clients who were asked to take HIVST to their sexual partners, with no differences in adverse events reported by arm. Our findings are similar to other secondary HIVST distribution strategies with both HIV-negative and status unknown women in Kenya and Malawi,^21-23^ even though ART clients (particularly women) potentially face greater risks due to HIVST distribution, such as status disclosure. High acceptability may have been due to the fact that ART clients in our trial were on ART for a mean of 5 years and over 90% had already disclosed to their partner prior to study enrollment. The majority of relationships were also highly stable – most were married, had children together, and were in a relationship for a mean of 9 years – which may result in greater trust and confidence in their relationship, facilitating safe HIVST distribution. Additional research is needed to assess how index HIVST performs among individuals in non-married and/or unstable relationships, particularly when the ART client has not yet disclosed their status.

Eight percent of index partners who used HIVST were unable to interpret their test result. Understanding the usability of HIVST within index testing is critical to ensuring the strategy is effective and scalable in low-resource settings, particularly where health literacy is low. The limited data available on the usability of secondary HIVST show that most partners are able to complete a HIVST test successfully, but some require guidance, desire pre-test counseling, or mistrust HIVST results.^24,37^ Additional research is needed to identify strategies that facilitate accurate test interpretation within secondary distribution models, such as additional visual aids and/or wider community sensitization about HIVST.

This is one of the first studies to examine ART initiation among HIVST users longitudinally. Twelve-months after enrollment, we found that only 47% of index partners who had a reactive HIVST kit initiated ART, and the vast majority of these individuals did so within the first 3-months after testing. Our experience is similar to other published data on secondary HIVST distribution, where linkage and initiation rates range from 23-68%.^25-28^ There are several potential explanations for low initiation rates within HIVST strategies. First, HIVST users may already be on ART but re-test because HIVST is convenient and does not require further conversations with providers, therefore removing the need for ART initiation. Although we are unable to confirm this hypothesis, other research in the region shows those with a known HIV status do retest for numerous reasons.^38^ Second, among those who did not previously know their status (or were aware but not on ART), simply knowing one’s status may not be enough to overcome traditional barriers to HIV service utilization (such as stigma, time and money required for facility visits, fear of unwanted disclosure at facilities, and fear of rude providers).^39^ Additional interventions could be coupled with index HIVST to address these ongoing barriers to care. Outside facility services, private and fast services, strong Welcome Back strategies, and peer counseling,^40,41^ may be required in order to optimize the impact of index HIVST.

This study has several limitations. First, ART clients who did not complete a four-week follow-up survey were excluded from the analyses because it was impossible to determine primary and secondary outcomes, which may result in bias if those retained were more likely to have used and be satisfied with the intervention. Second, we rely on secondary reports from ART clients to determine primary outcomes for their sexual partners. Secondary reports are likely to underestimate the impact of index HIVST since partners may use an HIVST kit, or receive additional HIV services, without the ART client’s knowledge. Third, our primary outcome (HIV testing) was measured 4-weeks after enrolment of the ART client, allowing only a short period for outcome attainment. Despite this limitation, we found high rates of testing in the index HIVST arm, and similar rates of testing in the PRS arm as compared to other studies. Finally, our sample size is small and the vast majority of ART clients were married and had already disclosed their HIV status to their partner. Additional research is needed to assess if findings are replicable in other facility types and other regions. Due to the small sample size, we had limited power to detect statistical differences in secondary outcomes.

## Conclusion

Index HIVST greatly increased HIV testing among partners of ART clients without increasing adverse events, however ART initiation after using HIVST remains a challenge. The intervention was cost-effective for new diagnoses, but improved ART initition is required to make index HIVST cost-effecitve for new ART initiations. Further research is needed to understand the role of index HIVST among non-stable partners, and what strategies can optimize testing among male partners and ART initiation more broadly.

## Data Availability

De-identified human data are available upon request from lead author [Dovel].

## Authors Contributions

KD and RH conceptualized the study. RH is responsible for funding acquisition. KD, RH, KB, FS, OA, BN, MN, VW developed study protocol and materials. KD, KB, KP, FS, EL, OO implemented the study. KD, KB, RH, SKG developed the analysis plan and KD, KB, EL analyzed the data. KD wrote the first draft and KB, KP, FS, OO, SKG, VW, EL, BN, MN, TM, AW, RH edited following drafts. All authors have read and approved the final manuscript.

## Acknowledgements

The authors are grateful to the research assistants who contributed to data collection, as well as study participants and health facility staff for their dedicated contribution to the study. The project was supported by the U.S. Agency for International Development (USAID) and the President’s Emergency Plan for AIDS Relief (PEPFAR) under Cooperative Agreement AID-OAA-A-15-00070. OO’s time was supported by the Fogarty International Center of the National Institutes of Health (NIH) under Award Number D43TW009343 and the University of California Global Health Institute (UCGHI). KD’s time was partially funded by the National Institute of Mental Health (NIMH) through T32MH080634-10. KD and RH receive support from the UCLA CFAR grant AI028697 and the UCLA AIDS Institute.

## Disclaimer

The findings and conclusions of this article are those of the authors and do not necessarily represent the official position of USAID or the US Government or other agencies represented.

